# Causal Emergence of Macroscopic Stochastic Dynamics from a Multi-modal Foundation Model of Human Sleep

**DOI:** 10.64898/2026.11.25.26368940

**Authors:** Yiwei Liu, Guangmeng Nie, Yu Sun, Hong Zhang, Junbo Zhang, Zhipeng Wang, Mingzhe Yang, Jingying Ye, Dong Li

**Affiliations:** Department of Pharmacy, Beijing Tsinghua Changgung Hospital, School of Clinical Medicine, Tsinghua Medicine, Tsinghua University; Sleep Medicine Center, Beijing Tsinghua Changgung Hospital, School of Clinical Medicine, Tsinghua Medicine, Tsinghua University; School of Systems Science, Beijing Normal University; Medical data science center, Beijing Tsinghua Changgung Hospital, School of Clinical Medicine, Tsinghua Medicine, Tsinghua University

## Abstract

Sleep is an essential, self organizing physiological state conserved across species, yet its macroscopic coordination remains governed by unknown physical principles. For nearly sixty years, clinical medicine has classified sleep into heuristic, discrete 30 second epochs, an empirical convention that flattens continuous non-equilibrium dynamics and leaves deep subcortical neuromodulatory control fundamentally unobservable. Although emerging artificial intelligence foundation models can decode multimodal physiology with high accuracy, their high-dimensional representations remain uninterpretable black boxes, leaving the generative laws of sleep unformulated. Here we show that, across four independent multinational cohorts, causal emergence analysis of a multimodal foundation model condenses human sleep into a single continuous order parameter whose dynamics are well described by a Landau Ginzburg jump diffusion equation.

This trajectory is physiologically structured: its overnight variance follows an AASM stage hierarchy (Wake, NREM, REM), and its diffusion shape tracks apnea hypopnea severity. Bidirectional clinical perturbations further support the decomposition: continuous positive airway pressure lowers baseline diffusion and apnea band spectral excess while shallowing the late-night potential well, whereas GABAergic agonism reduces the true jump rate and nocturnal awakenings without deepening the well. Length matched comparisons under dexmedetomidine sedation deepen the well and suppress jumps relative to natural sleep. By replacing empirical classification with an explicit stochastic description, our work uses a foundation model as a probe of physiological dynamics rather than only as a predictor and treats human sleep as an analytically tractable physical system. More broadly, the finding that multi-organ coordination during sleep can be summarized by a low dimensional order parameter suggests a general route from opaque artificial intelligence representations to testable macroscopic models of the subcortical dynamics that sustain human consciousness and fail in neuropsychiatric disease.

Graphic Abstract
A unified Landau–Ginzburg framework of human sleep dynamics.
Multimodal polysomnography (PSG)—EEG, respiratory effort (RESP), electrocardiogram (ECG), and electromyogram (EMG)—is discretized into 5-s tokens and encoded by a pretrained multimodal transformer (SleepFM) with a 5-min receptive context into a 512-dimensional embedding space. A shared PCA baseline () provides a microscopic reference, and the Neural Information Squeezer Plus (NIS+) information bottleneck evaluates causal emergence. Across cohorts, effective information is maximized at, defining a continuous one-dimensional order parameter that tracks the macroscopic sleep manifold over the night.

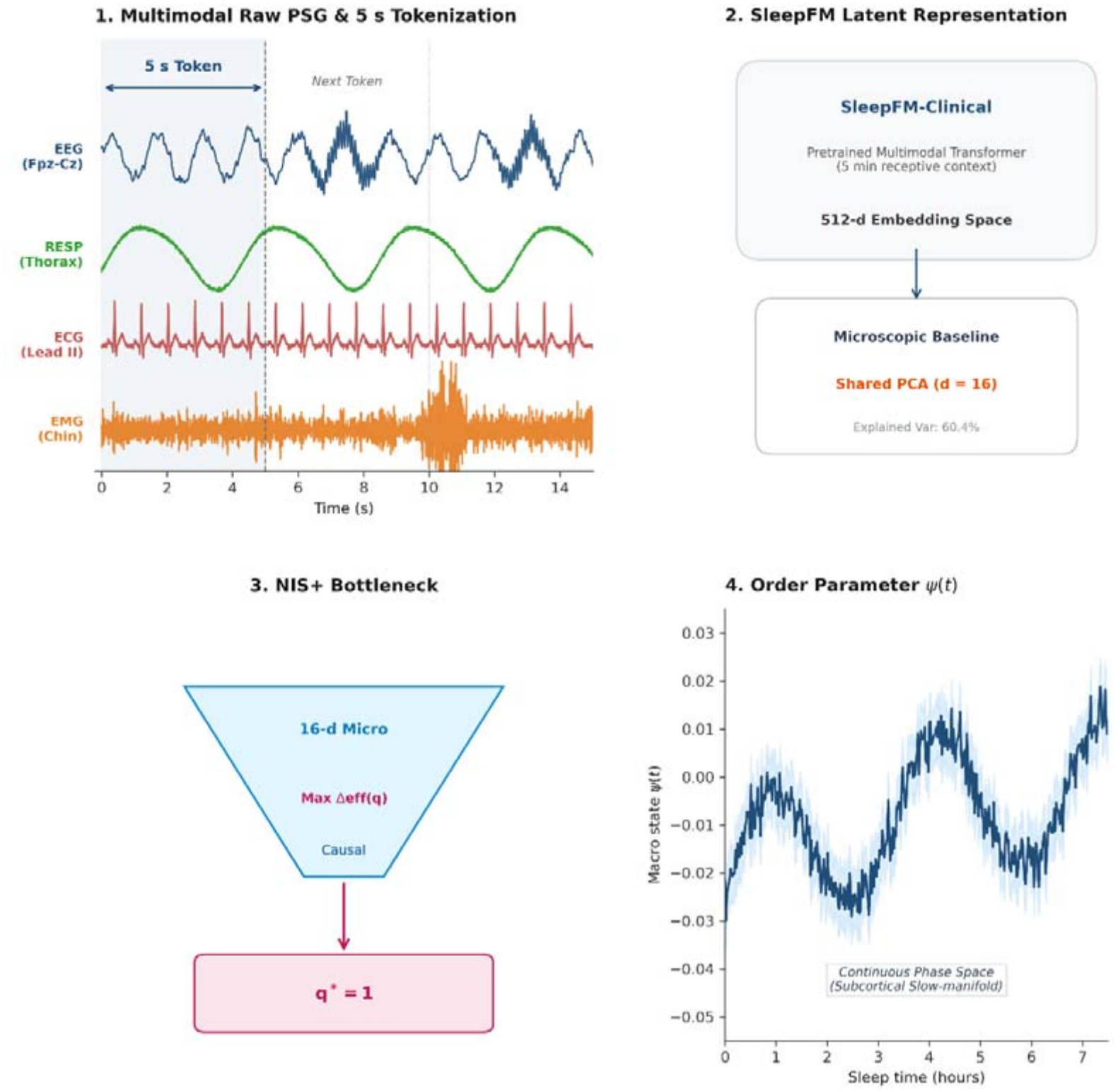

## Introduction

Far from being a passive suspension of consciousness, mammalian sleep operates as an active, non-equilibrium dynamical process—perpetually balancing homeostatic programming against intrinsic and environmental perturbations[1]. In this critical regime, an imperceptible subcortical event can trigger macroscopic cascades that ripple across the entire autonomic nervous system, reflecting the shifting topography of underlying attractors[2]. Yet, for more than half a century, clinical medicine and neuroscience have reduced this continuous phase-space journey to an epoch-averaged sequence of discrete 30-second stages—a legacy of 1960s paper-chart recordings that flattens complex non-linear dynamics into static boxes[3]. While pioneering frameworks of sleep microstructure have long argued that the sleeping brain does not operate in arbitrary clockwork slices[4], formulating an explicit, data-driven physical law that governs these continuous transitions directly from human multimodality has remained fundamentally out of reach.

To date, a profound divide has separated the microscopic components of sleep from its macroscopic clinical phenotypes[5]. At the microscopic extreme, cellular neurobiology has mapped an intricate web of subcortical arousal nuclei, neuromodulatory tides, and local cortical assemblies. At the macroscopic extreme, clinical polysomnography relies on coarse phenomenological metrics such as the apnea–hypopnea index (AHI) or sleep efficiency, which offer limited insight into underlying dynamical stability or therapeutic reversibility. To bridge this multiscale divide, mesoscopic theories of neural self-organization—most notably stochastic Landau–Ginzburg field formulations[6]—have demonstrated that collective cortical states naturally organize into distinct phases, including bistable up/down attractors characteristic of deep sleep and oscillatory regimes at the edge of synchronization. Yet, translating these field-theoretic principles to living human physiology has faced a formidable empirical barrier: real-world clinical recordings comprise a noisy, non-stationary mixture of neuroelectric, respiratory, and autonomic signals that historically lacked a principled coordinate mapping to theoretical order parameters. Consequently, the empirical realization and continuous tracking of an effective sleep dynamical theory directly from human data have remained out of reach.

Here, we test whether modern multimodal foundation models, combined with information-theoretic causal emergence, can provide the lens required to resolve this dilemma. Foundation models pre-trained on large collections of clinical bio-signals, such as SleepFM[5], do not merely act as automated stage classifiers; through cross-modal contrastive learning, they compress high-dimensional physiological streams into a representation that may reflect the upstream central coordinators of sleep. By applying the Neural Information Squeezer Plus (NIS+) [7] to this representation, we test whether the multi-organ activity of human sleep collapses onto an unobserved, continuous macroscopic manifold, and whether its dynamics can be written as an explicit stochastic equation. Foundational theories of brain dynamics propose that macroscopic integration emerges from a low-dimensional slow manifold, but direct quantitative validation in the awake, input-driven brain is difficult. Sleep, with continuous polysomnography and well-defined interventions, offers a more closed setting in which an effective equation can be derived directly from human data and a ‘black box’ transformer model, and then challenged with clinical perturbations.

## Results

### Multimodal causal emergence reveals a universal one-dimensional order parameter

To determine whether the complex, multi-organ dynamics of human sleep can be compressed beyond heuristic discrete staging, we developed an unsupervised causal emergence framework using the foundation model SleepFM (**Graphic Abstract**). Baseline characteristics of all cohorts are shown in **Supplementary Table S1**. Continuous polysomnography (PSG) encompassing four physiological streams—electroencephalogram (EEG), respiratory effort (RESP), electrocardiogram (ECG), and electromyogram (EMG)—was discretized into 5-s tokens with a 5-min receptive context, projecting the raw physiological dynamics into a 512-dimensional embedding space. To establish a microscopic reference while mitigating high-dimensional covariance, embeddings were reduced to a shared *d* = 16 baseline (capturing 60.4% of total quantify the gain in effective information Δeff(q) = eff(q) – eff(16) across macroscopic variance). We then applied NIS+ to evaluate causal emergence, using an information bottleneck to quantify the gain in effective information Δeff(*q*) = eff(*q*) − eff(*16*) across macroscopic dimensions *q* ∈ [1,16] (**Fig. 1A**).

**Figure 1.**
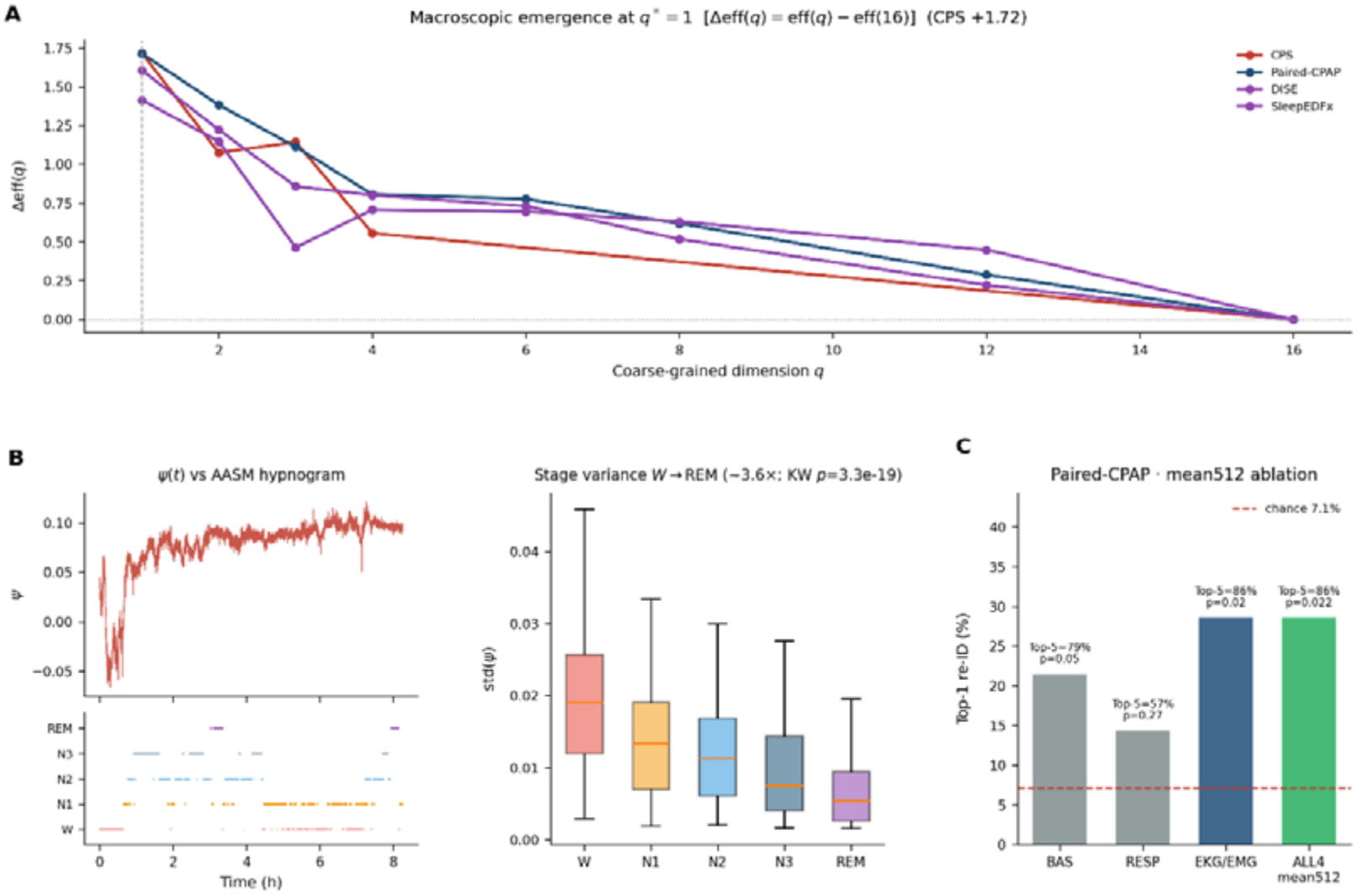
Causal emergence identifies a universal one-dimensional sleep macro-state across four independent cohorts. **(A)**Net causal emergence across coarse-grained dimensions estimated via Neural Information Squeezer Plus (NIS+) on sliding-window center-token SleepFM-Clinical embeddings (300-s context, 5-s stride, fixed center position index = 30). All four cohorts—CPS, Paired-CPAP, DISE and Sleep-EDFx—consistently peak at a one-dimensional macro-state. Horizontal dashed line denotes. **(B)**Left: Continuous 1D trajectory (top) aligned with the discrete 30-s AASM hypnogram (bottom) for a representative CPS diagnostic night, illustrating large-amplitude excursions during wakefulness and progressive stabilization across deeper sleep stages. Right: Within-stage fluctuation amplitude across CPS nights, revealing a monotonic ~3.6-fold reduction from wakefulness (W) to REM sleep (two-sided Kruskal–Wallis test,). Center lines indicate medians, boxes span the interquartile range (IQR), and whiskers extend to (outliers omitted for clarity). **(C)**Cross-night (diagnostic to CPAP) residual individual re-identification in the Paired-CPAP cohort using night-averaged 512-dimensional embeddings (mean512) after regressing out stage composition, apnea severity, and demographics. Bars show Top-1 accuracy for single-modality subsets (BAS, RESP, EKG/EMG) and four-modality fusion (ALL4 mean512), with Top-5 accuracies and permutation-values annotated above each bar (EKG/EMG and ALL4 mean512 both achieve 28.6% Top-1 and 86% Top-5 accuracy, and; red dashed line indicates 7.1% random chance).

Across four independent, multinational, and cross-hardware cohorts (Paired-CPAP; CPS; DISE; SleepEDFx), causal emergence profiles showed a single peak at q*=1 (**Fig. 1A**). In all cohorts, Δeff(q) declined monotonically for q>1 (CPS Δeff(1)=+1.72; Paired-CPAP +1.71; DISE +1.41; SleepEDFx +1.61), indicating that the cross-modal macroscopic causal structure of whole-night sleep is best summarized by a one-dimensional continuous order parameter, denoted *ψ*. The continuous order parameter *ψ* also captured dynamics that are not visible in conventional AASM staging (**Fig. 1B**). Night-level stage-wise standard deviation of *ψ* formed a clear hierarchy Wake > N1 > N2 > N3 > REM (~3.6× Wake-to-REM; Kruskal–Wallis p=3.3×10□^1^□), and continuous *ψ* (t) trajectories tracked the overnight hypnogram without being reducible to discrete 30-s stage labels.

This low-dimensional manifold was not recovered from isolated modalities. In a cross-night patient re-identification task on Paired-CPAP nights using mean512 embeddings **(Fig. 1C**), BAS-only and RESP-only Top-1 accuracies were 21.4% (Top-5=79%, permutation p=0.050) and 14.3% (Top-5=57%, p=0.27), respectively (chance=7.1%). EKG/EMG and four-modality both reached Top-1=28.6% with Top-5=86% (p=0.02), indicating that the macroscopic state *ψ* benefits from multi-organ coupling rather than localized cortical rhythms alone.

### Potential-well dynamics and clinical anchoring of diffusion shape

To interrogate the physical topography governing *ψ* (*t*), we performed non-parametric Kramers–Moyal (KM) stochastic inversion to reconstruct the empirical potential landscape (**Fig. 2A**):

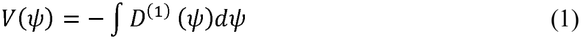

**Figure 2.**
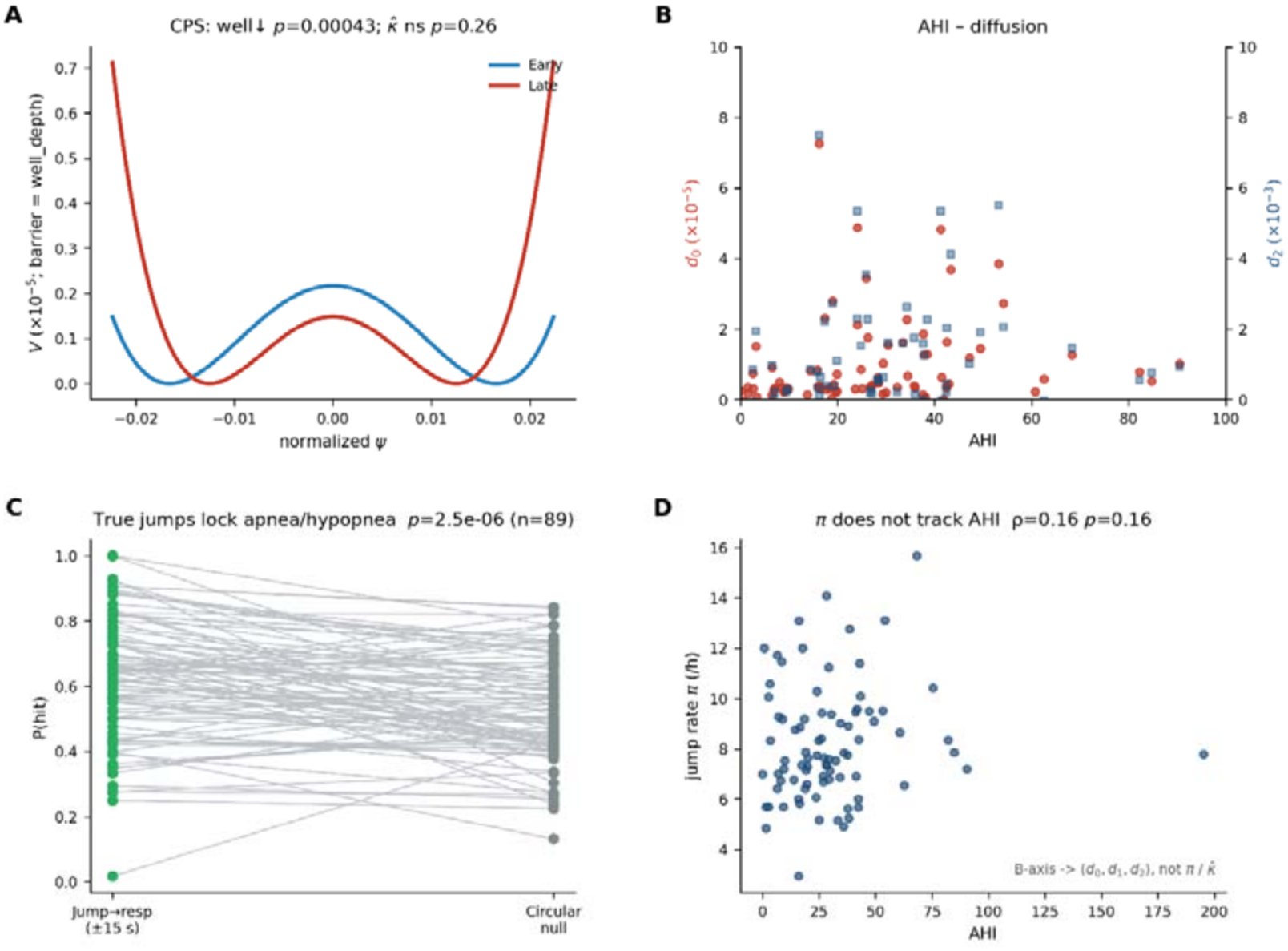
Overnight potential-well unloading and clinical anchoring of continuous diffusion and discrete jumps on the clean center-token manifold (CPS cohort). **(A)** Cohort-averaged symmetric quartic potential landscapes reconstructed from the early (first 3.5 h, blue) and late (last 3.5 h, red) halves of diagnostic nights, vertically shifted so that each well minimum equals zero (barrier = well_depth). Overnight homeostatic unloading significantly reduces well_depth (early vs. late paired test,; phase-randomized surrogate null,), whereas local bottom curvature remains unchanged (). **(B)** Cross-sectional association between apnea–hypopnea index (AHI) and quadratic multiplicative diffusion coefficients (red circles, left axis) and (blue squares, right axis) from (: Spearman,; :,). Higher OSA severity scales with elevated state-dependent diffusion turbulence rather than potential-well curvature. **(C)** Event-locked alignment between whitened Poisson jumps (,) and scored apnea/hypopnea events within a window. Observed hit probability (green) significantly exceeds within-night circular-shift null expectations (grey; paired Wilcoxon). **(D)** Scatter plot of hourly whitened jump rate () against AHI showing no significant association (Spearman,), consistent with adaptive whitening by absorbing continuous OSA severity into the diffusion axis () rather than or.

In the CPS diagnostic cohort (n=92 nights; paired early/late windows), comparing the first and second halves of the night revealed selective deformation of the potential well: well depth decreased late versus early (paired p=6.8×10□ □), whereas local curvature κ□ showed no reliable late–early change (p=0.26). Phase-randomized surrogates did not reproduce the well-depth shift (p=0.34), confirming that overnight unloading depends on non-equilibrium temporal structure rather than the linear power spectrum alone (**Fig. 2A**). To parameterize this landscape parsimoniously, the empirical potential was approximated by a minimal quartic Landau–Ginzburg (LG) normal form:

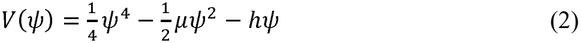

where µ governs barrier stability and *h* denotes a linear tilt field. Clinical OSA severity mapped specifically onto the state-dependent multiplicative diffusion coefficients of *ψ* (**Fig. 2B**): *d*_0_, *d*_1_, *d*_2_ correlated significantly with AHI (*ρ* =+, −0.33, and + 0.32; *p* =0.0027, and 0.0035). Higher AHI therefore corresponds to elevated baseline turbulence, a more negative linear tilt, and stronger U-shaped diffusion divergence away from the attractor core, rather than to stiffening of the local potential minimum. Finally, discrete jumps on the clean manifold locked selectively onto respiratory events rather than cortical arousals or macro-stage switches (**Fig. 2C–D**). In CPS recordings with scored event annotations, the probability that a whitened jump (|*z*| > 3) coincided within ±15 s of a scored apnea/hypopnea exceeded within-night circular-shift nulls (paired Wilcoxon *p* = 2.5× 10 −6), whereas alignments with cortical EEG arousals (*p* = 0.82) and AASM stage transitions (*p* = 0.29) were nonsignificant. Whitened jump intensity π itself did not track AHI (*ρ* = 0.16, *p* = 0.16; **Fig. 2D**), consistent with adaptive normalization by 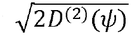 absorbing continuous OSA severity into the diffusion axis (*d*_0_,*d*_1_,*d*_2_) while discrete jumps capture acute respiratory-coupled shocks.

### Empirical identification of a stationary jump-diffusion stochastic differential equation

In non-equilibrium statistical mechanics, coarse-graining fast microscopic degrees of freedom generically gives rise to effective Langevin dynamics governed by state-dependent, multiplicative noise rather than simple additive fluctuations [8]. Consistent with this principle, empirical evaluation of the conditional second moment in the clean, chunk-clock-free macroscopic sleep manifold, *ψ*(*t*) revealed systematic state dependence in both fluctuation amplitude and intermittent jumps. We therefore sought the minimal, sufficient, and unconstrained stochastic differential equation (SDE) capable of generating the one-dimensional order parameter, *ψ*(*t*) intermittent jumps. We therefore sought the minimal, sufficient, and unconstrained stochastic within each analysis window, modeled as a jump–diffusion LG system:

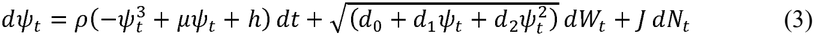

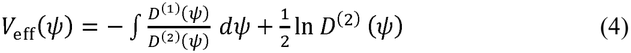

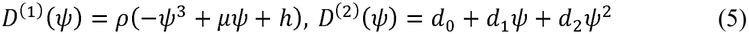

where *dW*_*t*_ is a standard Wiener increment, *dN*_*t*_ is a homogeneous Poisson counting process with intensity π, and *J* represents discrete jump amplitudes. The full-night recordings from the CPS cohort (sliding-window center-token embeddings), each component of Eq. (3) was sequentially tested and validated (**Fig. 3**). Evaluation of the empirical drift field *D*^(1)^ (*ψ*) under cross-subject leave-one-out (LOO) out-of-sample negative log-likelihood (NLL) confirmed that the cubic restoring force of the LG potential outperformed a linear Ornstein–Uhlenbeck (OU) relaxation baseline across clean center-token CPS recordings (**Supplemental Table S2**: LG outperformed OU in 73.0% of nights meeting LOO drift-estimation criteria, median ΔNLL_OU−LG_ = +0.0012 per 5-s step, paired Wilcoxon *p* = 3.3 × 10^−s^). Notably, because homeostatic well_depth systematically shallows from early to late night, intra-night temporal splits (early↔late) shift the outer-tail restoring force and no longer consistently favor the static cubic tail over linear OU; cross-subject LOO therefore provides the primary out-of-sample test of the shared cubic functional form, with the modest step-wise ΔNLL reflecting that cubic nonlinearity acts mainly during outer-flank excursions beyond the nearly linear well core. Because the linear OU drift is nested within the cubic polynomial, an in-sample gain is expected; this comparison establishes the adequacy of the cubic normal form rather than its uniqueness and motivates out-of-sample cross-validation below. Analysis of the conditional second moment *D*^(2)^ (*ψ*) rejected pure additive white noise in favor of state-dependent quadratic diffusion (**Fig. 3A–B**). Across clean CPS nights, unconstrained quadratic fits yielded a positive median curvature *d*_2_ = 3.43 × 10^−4^ (*d*_2_ >0 in 65.2% of nights), negative linear tilt *d*_1_ = −5.74 × 10^−5^ (*d* _0_ < 0 in 80.4% of nights), and joint U-shaped geometry (*d*_2_ > 0,*d*_1_ >0) in 64.1% of nights (*d*_0_ = 4.30 × 10^−6^; **Fig. 3A–B**)—a substantial and physiologically consistent pattern, though not universal across every individual recording. Notably, the diffusion minimum of the median quadratic curve (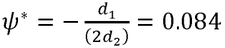; **Fig. 3B**) aligns closely with the sleep-stage attractor centers (median *ψ* = 0.077−0.086), whereas wider wake/arousal excursions populate the rising flanks. Importantly, under dexmedetomidine sedation (DISE cohort), projecting recordings onto the frozen CPS center-token encoder and length-matching CPS to 50-min segments abolished any apparent diffusion-shape reversal: short-window *d*_2_ estimates were highly variable in both cohorts, but their distributions overlapped, *d*_2_ in 54% of DISE vs. 48% of CPS (Mann–Whitney p = 0.83; as did baseline diffusion *d*_0_ *p* = 0.94). Instead, relative to length-matched natural sleep, dexmedetomidine clamped the trajectory into a markedly deeper and stiffer confining well (well depth: 1.23 × 10^−5^ vs 9.60 × 10^−7^, *p* = 7.1 × 10^−31^;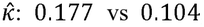, *p* = 7.4 × 10^−12^; **Fig. 3C**) with suppressed jump intensity (*π*: 0.00749 vs 0.01169, i.e., 5.4 vs 8.4 events h^−1^, *p* = 3.0 × 10^−4^; **Fig. 3D**). Notably, the apparent reduction in overall dispersion *y*_std_ relative to full-night CPS (0.0115 vs 0.0155) proved to be a recording-duration artifact, as *y*_std_ under DISE was in fact higher than in length-matched 50-min CPS segments (0.0115 vs 0.00855, *p* = 4.1 × 10^−7^; **Fig. 3D**).

**Figure 3.**
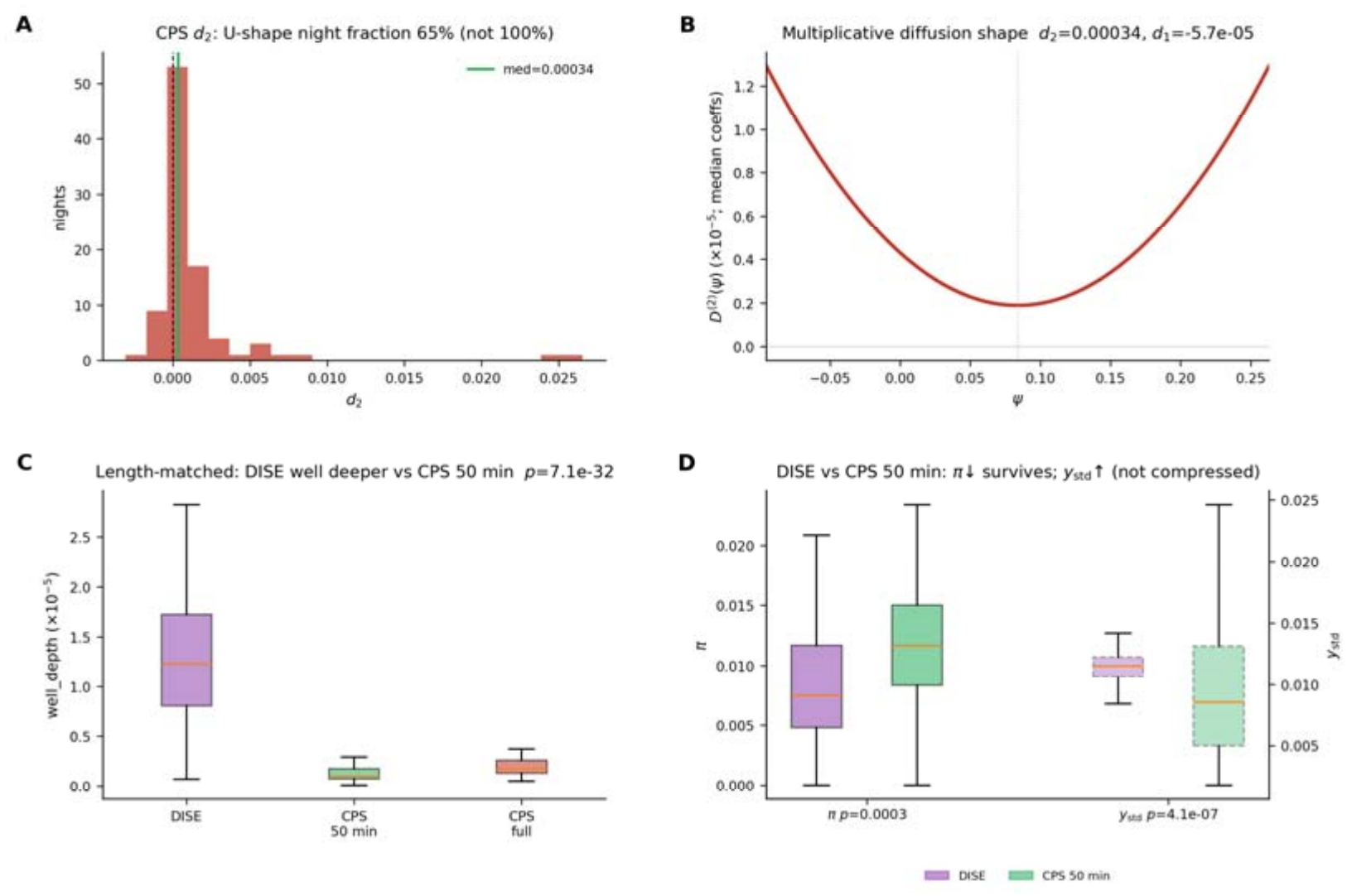
Multiplicative diffusion geometry on the purified manifold and length-matched DISE contrast. All analyses use the unified sliding-window center-token SleepFM manifold projected onto the frozen German CPS encoder (*q*^*^ = 1). **(A)** Distribution of the quadratic diffusion coefficient *d*_2_ across CPS nights (*d*_2_ = 0; solid green line, cohort median 3.43 × 10^−4^). Positive curvature (*d*_2_ > 0) occurs in 65.2% of nights (64.1% full U-shape with *d*_2_ > 0, *d*_1_ < 0). **(B)** Representative multiplicative diffusion profile *D*^(2)^(*ψ*) = *d*_0_ + *d*_1_*ψ* + *d*_2_*ψ*^2^ evaluated at CPS cohort median coefficients (*d*_0_ = 4.30 × 10^−6^, *d*_1_ = −5.74 × 10^−5^, *d*_2_ = 3.43 × 10^−4^). **(C)** Length-matched potential well depth (well_depth) comparing dexmedetomidine-induced sleep endoscopy (DISE, median duration 40 min) against length-matched 50-min CPS segments (median of three random 600-token windows per night; two-sided Mann–Whitney *U* test, *p* = 7.1 × 10^−32^); full-night CPS is shown for reference. **(D)** Length-matched jump rate *π* (left axis, solid boxes; DISE vs. CPS 50-min, *p* = 3.0 × 10^−4^) and trajectory dispersion *y*_std_ (right axis, dashed boxes; DISE vs. CPS 50-min, *p* = 4.1 × 10^−7^). Relative to duration-matched natural sleep, dexmedetomidine anesthesia locks the system into a deeper well with fewer discrete jumps while exhibiting slightly greater within-window dispersion rather than amplitude compression. Boxplots show median (orange line), interquartile range (box), and 1.5 × interquartile range (whiskers).

Even after accounting for multiplicative diffusion, whitened residuals retained excess kurtosis, revealing intermittent non-Gaussian jumps. On the clean center-token manifold, median jump intensity was π = 0.0106 (~ 7.6 events h^−1^)—approximately one-third of the ~ 20–24 h^−1^ rate inflated by 5-min chunk-boundary discontinuities in naive block embeddings—and remaining physiological jumps were significantly enriched around respiratory events (observed hit rate 0.64 vs null 0.53, *p* = 2.5 × 10^−6^) rather than cortical EEG arousals or stage transitions.

Cross-subject LOO out-of-sample NLL evaluation across the stochastic model hierarchy (**Supplemental Material Table S2**) confirmed that each stochastic term contributes genuine predictive structure rather than in-sample overfitting. Specifically, quadratic multiplicative Gaussian diffusion outperformed additive noise in 85.4% of nights (median ΔNLL = +0.024 per 5-s step, Wilcoxon *p* = 3.4 × 10^−8^). Replacing Gaussian multiplicative residuals with heavy-tailed Student-t noise yielded a modest further gain in 55.1% of nights (median ΔNLL = +0.0077, 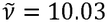, *p* = 2.3 × 10^−4^). Crucially, however, explicitly incorporating discrete Poisson jumps decisively outperformed both continuous-noise baselines in 100% of nights by a large margin (jump–diffusion vs multiplicative Gaussian: median ΔNLL = +2.39, *p* = 1.3 × 10^−16^; jump–diffusion vs multiplicative Student-*t*: median ΔNLL = +2.37, *p* = 1.3 × 10^−16^).

Finally, we verified global generative closure of the full SDE against empirical whole-night occupancy – ln 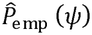. In the Itô/Risken convention, the pure-diffusion (*J* = 0) stationary effective potential Eq. (4) accurately recovers simulated log-densities (median *R*^2^ = 0.990, compared with 0.854 when the entropic term is misspecified as 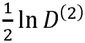, whereas introducing Poisson jumps *JdN*_*t*_ shifts the stationary density away from the pure-diffusion Boltzmann limit (R^2^ = 0.780, Δ = −0.21, p = 10^−15^). Crucially, forward simulation of the full jump–diffusion SDE (Eq. 3) on clean center-token CPS trajectories reproduced the empirical whole-night log-density 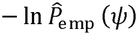 with a median *R*^2^ of 0.799, representing a dramatic improvement over naive non-overlapping block embeddings (*R*^2^ = 0.319, Wilcoxon *p* = 3.74 × 10^−15^). Together, these local out-of-sample likelihood and global generative density tests establish that *ψ*(*t*) obeys a stationary jump–diffusion LG law with multiplicative diffusion and respiratory-coupled jumps.

### Bidirectional in vivo perturbations: CPAP unloads diffusion load; temazepam suppresses true jumps

To test causal validity, we examined the model under bidirectional physical and pharmacological perturbations using a frozen CPS encoder so that paired nights share one physical ruler. First, in the Paired-CPAP cohort, therapeutic CPAP reduced baseline diffusion *d*_0_ (Wilcoxon p=0.025) and 30–60 s apnea-band spectral excess (p=0.004, **Fig. 4A**), while flattening the velocity spectral slope *α*_*v*_ (*p* = 0.002) and shallowing late-night well_depth on the same ruler (*p* = 0.003; **Fig. 4B**); κ□ did not respond (p=0.54). Second, in the double-blind temazepam crossover (SleepEDFx), the drug lowered the true jump rate (jump/h 5.63→4.71; *p*_one-sided_ = 4.9 × 10^−5^, *p*_two-sided_ = 9.9 × 10^−5^), the absolute unwhitened jump rate (11.29 → 8.53 h^−1;^ *p*_two-sided_ = 0.016), and clinically scored long awakenings (*NAW* ≥ 60 s; median Δ = −2.0, *p*_one-sided_ = 0.009, *p*_two-sided_ = 0.018; **Fig. 4C**), while steepening *αv* (*p* = 5.0 × 10^−4^). By contrast, continuous well_depth (*p* = 0.82), baseline diffusion *d*_0_ (*p* = 0.48), and apnea-band spectral excess were unaffected, and the group-level escape fraction (*NAW*/*n*_*jump*_) did not decrease (0.130 → 0.154, *p* = 0.17). Furthermore, within-subject changes in jump rate (Δ*π*) did not correlate linearly with Δ*NAW* (Spearman *ρ* = 0.09, *p* = 0.71; **Fig. 4D**), indicating that although temazepam consistently suppresses discrete attempt frequency at the cohort level, individual awakening counts remain modulated by subject-specific escape susceptibility. Framed within a Kramers escape perspective (*r*_*K*_ ∝ *ν*_0_*e*^−Δ*V*/*D*^), the clean center-token manifold therefore reverses the earlier block-artifact reading that attempt frequency *π* was invariant (~ 21 h^−1^) under temazepam: the drug acts primarily by suppressing discrete jump attempts (*ν*_0_ ~ *π*) rather than by deepening the continuous potential barrier (Δ*V*).

**Figure 4.**
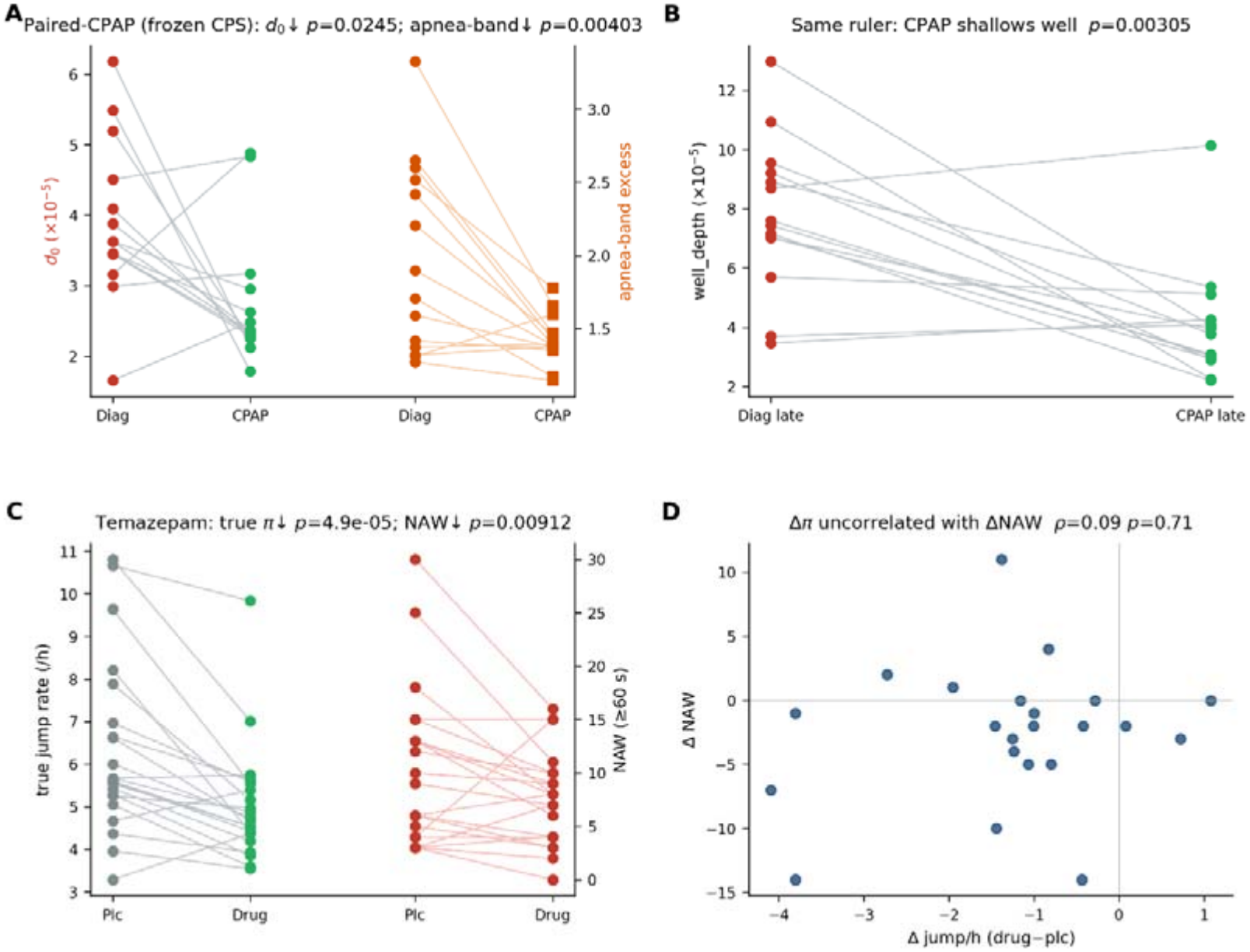
Dissociated mechanical and pharmacological perturbations on the frozen CPS center-token ruler. All trajectories were extracted via sliding-window center-token SleepFM embeddings and zero-shot projected onto the frozen CPS NIS+ coordinate. **(A)** Paired CPAP cohort: continuous baseline diffusion (left axis, red/green circles; two-sided paired Wilcoxon) and 30–60 s apnea-band spectral excess (right axis, orange circles/squares;) both decrease under CPAP. **(B)** Late-night potential well depth (last 3.5 h) significantly shallows under CPAP on the same frozen ruler (). **(C)** Sleep-EDFx temazepam crossover cohort (placebo vs. drug): pharmacological GABA-A modulation suppresses the true discrete Poisson jump rate (, left axis;-,-) and clinical awakenings lasting (, right axis;-,-). **(D)** Within-subject changes in jump rate (, drug minus placebo) and awakenings (, Spearman,), showing that while temazepam consistently reduces both attempt rate and awakenings at the cohort level (mostly negative quadrants), individual awakening reduction is not linearly proportional to.

#### Foundation-model-free cross-scale validation on raw EEG beta-envelope dynamics

To verify that the Landau–Ginzburg Kramers-escape structure is an intrinsic property of human sleep neurophysiology rather than an artifact of SleepFM or NIS+ compression, we applied the same stochastic inversion pipeline directly to raw cortical EEG beta-band power envelopes 15–30 Hz, ±5 s around micro-arousals without foundation-model embeddings or bottleneck training) across the CPS cohort (**Table 1**). Consistent with scale-invariant barrier-crossing kinetics at the cortical micro-arousal level, the effective potential barrier height Δ*V*_*β*_ estimated purely from the raw beta envelope correlated strongly and inversely with manual clinical ARI (Pearson *r* = −0.61, *p* = 10^−9^, *ρ* = −0.51), whereas the raw jump attempt rate alone did not significantly predict ARI (*r* = −0.21, *p* = 0.057; *ρ* = −0.049). Combining barrier height and attempt rate via the closed Kramers escape rate *P escape, β* = *c* · *πβ* exp(−Δ*Vβ*/*Dβ*) robustly tracked clinical ARI (*r* = +0.58, *p* = 10^−8;^ *ρ* = 0.45). Together, these model-free results confirm that the same Kramers jump-diffusion physics governs arousal transitions across both raw microscopic cortical envelopes and SleepFM’s macroscopic multi-organ order parameter *ψ*(*t*).

**Table 1.** Kramers escape kinetics against clinical micro-arousal index (CPS cohort, n=82)

| **Predictor** | **Physical Interpretation** | **Pearson *r* [95% CI] to ARI** | **P value** | *ρ* | **Hypothesis Verdict** |
|---|---|---|---|---|---|
| *π* | Intrinsic subcortical attempt rate | -0.21[-0.41, 0.01] | 0.057 | −0.049 | Not independently predictive |
| Δ*V* | Effective potential barrier height | -0.61 [-0.74, -0.45] | 10<sup>−9</sup> | −0.51 | Supported (barrier containment) |
| *P*<sub>escape</sub> | Kramers rate:<br>*c* · *π*exp (−Δ*V*/*D*) | 0.58[0.42, 0.71] | 10<sup>−8</sup> | +0.45 | Confirmed (closed kinetics) |

### Clinical anchoring, jump specificity, and length-matched anesthesia control

Finally, we evaluated the specificity and boundary conditions of the de-clocked dynamical model (Fig. 5). Within the German CPS cohort, Spearman rank correlations against AHI confirmed that OSA severity is anchored by the multiplicative diffusion coefficients (*d*_0_: *ρ* = 0.33, *p* = 0.0027; *d*_1_: *ρ* = −0.33, *p* = 0.0022; *d*_2_: *ρ* = 0.32, *p* = 0.0033), whereas local drift curvature 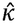 was nonsignificant (Fig. 5A). Event-alignment analysis further established that discrete jumps in *ψ* (median rate *π* = 0.0106) significantly lock onto respiratory events (*p* = 2.5 × 10^−6^), while showing no enrichment around cortical EEG arousals (*p* = 0.82) or macro-stage transitions (*p* = 0.29; **Fig. 5B**). Thus, overnight homeostatic unloading manifests in well_depth, whereas OSA burden and acute perturbations map specifically onto state-dependent diffusion and respiratory-locked jumps. To distinguish pharmacological sedation effects from recording-duration artifacts, we compared dexmedetomidine-induced sleep endoscopy DISE cohort against both whole-night CPS and duration-matched 50-min CPS segments on the frozen encoder (**Fig. 5C**). Controlling for segment length confirmed that sedation produces a substantially deeper (well_depth, *p* = 7.1 × 10^−32^) and steeper (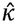, *p* = 7.4 × 10^−12^) confining potential with suppressed discrete jumps (*π, p* = 3.0 × 10^−4^) and slightly greater local dispersion (*y*_std_, *p* = 4.1 × 10^−7^), whereas multiplicative diffusion curvature (*d*_2_, *p* = 0.83) and baseline diffusion (*d*_0_, *p* = 0.94) remained indistinguishable from natural sleep. Notably, naïve comparison against whole-night CPS inverted the apparent direction of *y*_std_, underscoring the necessity of duration-matched baselines.

**Figure 5.**
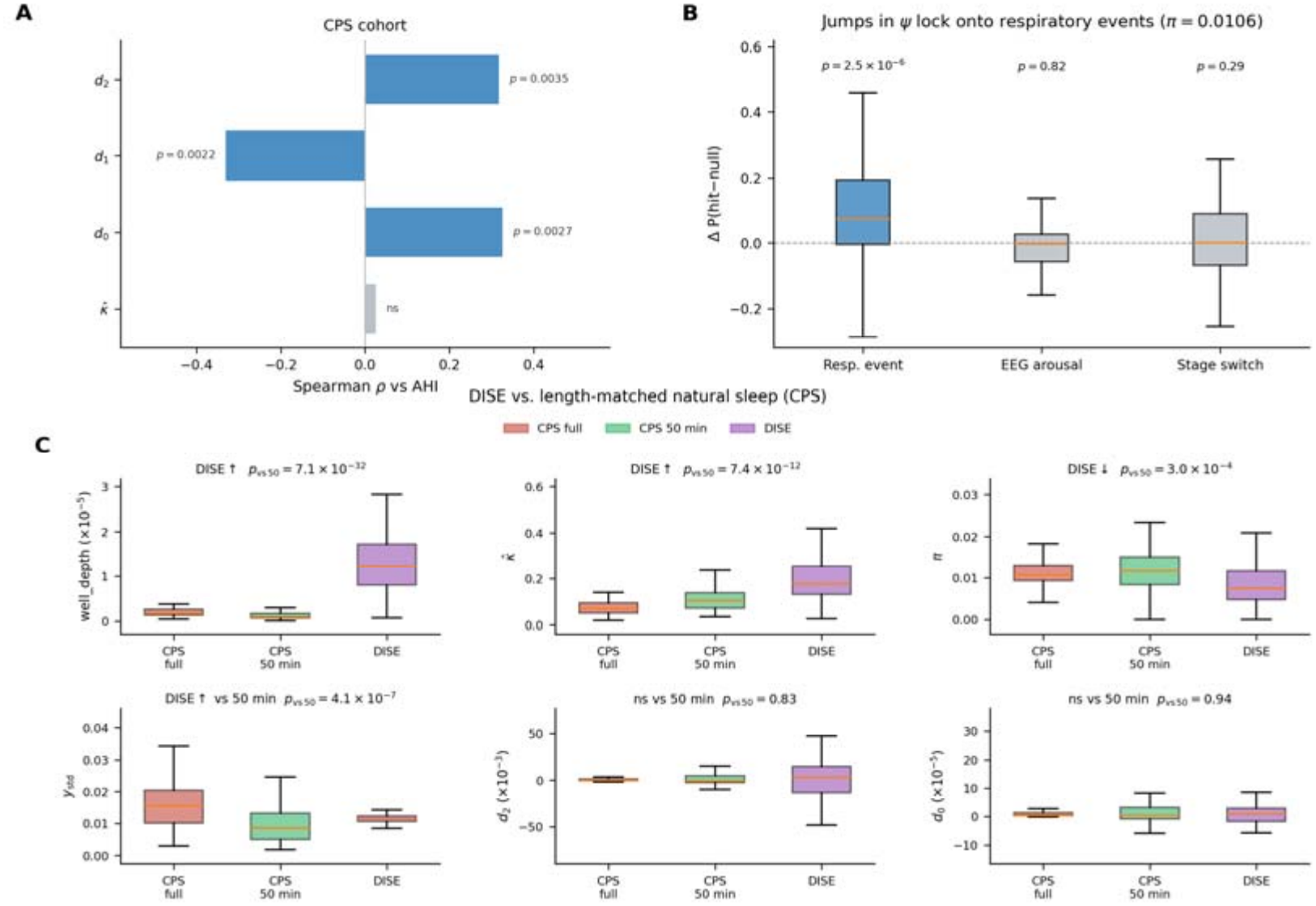
Clinical anchoring, jump specificity, and length-matched anesthesia control on the clean manifold. **(A)** Clinical severity anchoring in the German CPS cohort. Horizontal bars show Spearman rank correlation (*ρ*) between apnea–hypopnea index (AHI) and whole-night diffusion coefficients (*d*_0_, *d*_1_, *d*_2_) versus drift curvature 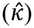. Multiplicative diffusion coefficients correlate significantly with AHI (*d*_0_: *p* = 0.0027; *d*_1_: *p* = 0.0022; *d*_2_: *p* = 0.0033; blue), whereas 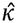 is nonsignificant (ns, grey). **(B)** Event-level physiological specificity of discrete jumps in *ψ* (median jump rate *π* = 0.0106 per 5 s step). Boxplots show the excess hit probability Δ*P*(hit − null) relative to a time-shuffled/uniform null (horizontal dashed line at 0). Jumps significantly lock onto respiratory events (*p* = 2.5 × 10^−6^, blue), but show no enrichment around cortical EEG arousals (*p* = 0.82) or macro-stage switches (*p* = 0.29, grey; paired Wilcoxon tests). **(C)** Dexmedetomidine-induced sleep endoscopy (DISE; purple) compared with whole-night natural sleep (CPS full, coral) and length-matched 50-min natural sleep segments (CPS 50 min, green, median across three random 600-token windows per night) on the frozen CPS encoder. Two-sided Mann–Whitney *U p*-values compare DISE against CPS 50 min (*p*_vs50_). After matching recording duration, DISE exhibits a substantially deeper (well_depth, *p* = 7.1 × 10^−32^) and stiffer (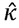, *p* = 7.4 × 10^−12^) confining potential with fewer discrete jumps (*π, p* = 3.0 × 10^−4^), despite slightly larger trajectory dispersion than 50-min natural sleep (*y*_std_, *p* = 4.1 × 10^−7^), while multiplicative diffusion curvature (*d*_2_, *p* = 0.83) and baseline diffusion (*d*_0_, *p* = 0.94) remain indistinguishable from natural sleep. Across all boxplots, center lines indicate medians, boxes span the interquartile range (IQR), and whiskers extend to 1.5 × IQR (3 outliers omitted for clarity).

For sensitivity analysis, the reconstructed the 1D manifold *ψ*(*t*) directly from single-channel peripheral pulse transit time (PTT) via Kalman filtering failed (late-night median *r* = 0.02, supporting the necessity of multimodal foundation embeddings. Gating drift parameters by instantaneous attractor occupancy Φ(*ψ*) explained negligible variance in Δ*μ* (*R*^2^ = 0.0082) and worsened model parsimony (ΔAIC = +224, favoring the ungated model), rejecting closed-loop micro-state feedback. When allowing an explicit exponential relaxation clock in *V*(*ψ, t*) yielded a physiologically plausible median timescale (*τ* = 4.0 h), but improved AIC in only 36% of nights (median ΔNLL = −0.0014, leaving the autonomous drift favored in 64% of nights). Thus, slow overnight homeostatic unloading is weak relative to 5-s step noise and is more robustly resolved through half-night potential contrasts than by adding an explicit within-night relaxation parameter.

## Discussion

The primary objective of this work was to determine which features of sleep neurophysiology are captured by high-capacity multimodal foundation models beyond classical heuristic scoring. Beyond this immediate goal, the analysis yielded a data-derived macroscopic order parameter for human sleep dynamics, obtained without imposing a predefined state variable; a Landau–Ginzburg phenomenology of sleep-stage transitions has recently been proposed[9]. This connects, in a limited and specific way, to a longstanding gap in the “easy problem” of consciousness. That problem is often treated as solved in principle, awaiting only empirical detail; in practice, however, it still lacks what classical statistical mechanics possesses: a rigorous macroscopic order parameter, a principled coarse-graining procedure, and an experimentally validated effective equation. We do not claim to resolve this gap. What we provide is one concrete instance—restricted to sleep, not to consciousness in general—in which all three ingredients are realized together: causal emergence supplies a data-driven coarse-graining step, the resulting one-dimensional manifold supplies an empirical order parameter, and Kramers–Moyal inversion supplies an empirically reconstructed effective equation whose parameters are interpretable, intervention-sensitive, and consistent with adiabatic elimination of microscopic fluctuations[10]. Whether the same order parameter and coarse-graining logic extend to wakeful consciousness remains an open question.

By combining SleepFM representation learning with NIS+ and Langevin inversion, we found in four natural-sleep cohorts, with consistent results in a sedation cohort, that human sleep macro-dynamics collapse onto a one-dimensional continuous order parameter, *ψ*(*t*) (*q*\* = 1) [11]. Rather than an arbitrary latent projection, this trajectory is well described by a Landau–Ginzburg stochastic differential equation with a cubic restoring force, state-dependent multiplicative diffusion, and asymmetric heavy-tailed jumps. This formulation helps bridge discrete phenotypic scoring and continuous biophysical dynamics and provides a quantitative framework for interpreting clinical and pharmacological perturbations. More generally, the approach may serve as a template for the mechanistic interpretation of foundation models in physiology.

### Theoretical Grounding of Transformer: From Modern Hopfield Free-Energy Minimization to Causal Macroscopic Emergence

A fundamental question arising from our findings is why a SleepFM trained purely on multi-modal contrastive objectives does not disperse representations across its nominal 512-dimensional embedding space, but instead naturally constrains physiological dynamics to an emergent, one-dimensional causal manifold (*q*^*^ = 1). From a machine learning perspective, this severe dimension collapse is often viewed as an empirical curiosity. However, through the lens of non-equilibrium statistical mechanics, this phenomenon admits a natural theoretical interpretation through the equivalence between scaled dot-product attention and modern continuous Hopfield networks. This connection provides an energy-based perspective on the organization of the learned representation, although it does not imply that the trained Transformer implements a literal thermodynamic equilibrium process.

The canonical scaled dot-product attention mechanism updates query tokens *Q* across key-value pairs (*K’V*) via:

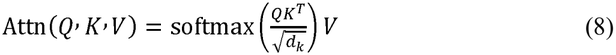

As formally established by modern continuous Hopfield network theory[12], this operation is mathematically identical to a single-step fixed-point minimization of a continuous energy functional. For a state vector 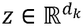 interacting with stored memory patterns *X* = [*x*_1_, …, *x*_*N*_], the global energy is defined by the Log-Sum-Exp potential:

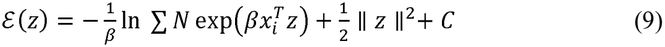

where the thermodynamic inverse temperature is precisely the architectural scale factor 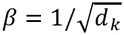. Under gradient descent on ε(*z*), the iterative update rule Δ*z* ∝ −∇*z* ε(*z*) yields the exact probability distribution:

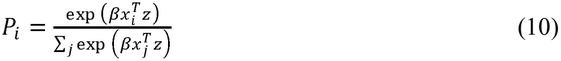

which is isomorphic to the classic Boltzmann–Gibbs equilibrium distribution, wherein the cross-modal dot-product 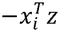 acts as the effective pairwise interaction energy. Consequently, as tokens representing EEG, EKG, EMG and RESP dynamics propagate through stacked Transformer layers, the network is not merely performing non-linear feature extraction; it is executing successive rounds of thermal relaxation on a high-dimensional free-energy landscape. In each layer, the high-entropy, uncorrelated fast fluctuations among peripheral physiological channels are progressively extinguished, pulling the composite state trajectory into the dominant metastable attractor basins shaped by normative human physiology.

While the internal representation resides nominally in a high-dimensional space, the macroscopic observational dynamics cannot sustain infinite degrees of freedom. In physical systems near critical transitions, renormalization-group arguments dictate that microscopic details wash out and the slowest collective mode dominates. From this perspective, the modern continuous Hopfield energy landscape explains why SleepFM’s 512-dimensional latent space has the structural capacity to organize multi-organ correlations into low-dimensional attractor basins rather than dispersing uniformly across 512 dimensions, enabling NIS+ to isolate a single dominant macroscopic coordinate (*q*\* = 1). Crucially, however, projecting a high-dimensional latent manifold down onto a one-dimensional scalar observable *ψ*(*t*) is an effective coarse-grained approximation rather than an exact copy of the full latent geometry. As in Mori–Zwanzig projection of unresolved degrees of freedom, the transverse dimensions eliminated by compression reappear in the 1D projected equation as state-dependent multiplicative diffusion *D*^(2)^(*ψ*) and intermittent non-equilibrium jumps *JdN*_*t*_—accounting for why the 1D empirical occupancy deviates from a pure constant-diffusion Boltzmann equilibrium (R^2^ = 0.78 vs. 0.99 when jumps are absent) even if the underlying high-dimensional representation is organized around energy-like basins. Within this effective 1D description, the reconstructed drift potential naturally takes the minimal quartic Landau–Ginzburg normal form permitted by symmetry for bistable homeostatic charging and state switching.

### Continuous Order-Parameter Dynamics versus Discrete Staging

For more than half a century, sleep medicine has relied on the 1968 Rechtschaffen and Kales criteria and its AASM revisions, which discretize continuous brain dynamics into 30-second epoch classifications. While pragmatic for manual scoring, this temporal and categorical discretization acts as a severe filter, obscuring sub-minute energy relaxations and critical fluctuations[3]. Our comparative analysis in the Paired CPAP cohort highlights this limitation: while the discrete AASM transition sequence was invariant across treatment nights, the continuous order parameter *ψ*(*t*) captured robust overnight variance relaxation. The collapse to *q*^*^ =1 was reproduced across diagnostic polysomnography, international validation cohorts, heterogeneous neurological cohorts, large-scale ambulatory recordings, and short-duration drug-induced sedation. This suggests that the collective activity of cortical, respiratory, cardiac, and somatosensory circuits is constrained to an invariant one-dimensional slow manifold during sleep.

### The Two-Axis Phenotypic Space and Therapeutic Dissociation

OSA severity instead mapped onto multiplicative diffusion shape (*d*_0_, *d*_1_, *d*_2_), while true jumps provide a respiratory-locked intermittent drive. Our perturbation analyses suggest that clinical interventions dissociate these coordinates. Acute CPAP reduces *d*_0_/apnea-band load and shallows the late well without changing κ□, indicating mechanical unloading of acute ventilatory work. Temazepam reduces true jump rate and NAW without deepening the well, arguing against a simple barrier-height Kramers account on the clean manifold. DISE deepens the well and suppresses jumps relative to length-matched natural sleep without flipping *d*_2_. Our formulation focuses on non-equilibrium relaxation dynamics within the nocturnal sleep window. Within this timeframe, the 24-hour circadian drive (Process C) acts approximately as an adiabatic reservoir, consistent with our finding that an explicit within-night relaxation clock did not improve the fit. Capturing the complete 24-hour cycle would require continuous multi-day recordings; this is outside the scope of the present study, which aims to identify the minimal stochastic differential equation describing within-sleep dynamics and clinical state transitions.

### Pharmacological intervention reshapes the effective potential: Kramers escape and distinct fluctuation regimes

The two interventions therefore probe different aspects of the same potential. CPAP selectively unloads respiratory/turbulence load (*d*_0_↓, apnea-band↓, late well↓). Temazepam suppresses jump attempts (π↓) and long awakenings without a detectable well-deepening effect. Dexmedetomidine sedation (DISE) deepens/steepens the landscape and lowers π versus length-matched CPS, without the previously claimed inverted-*d*_2_ geometry once encoder and duration are matched. Across interventions, microscopic jump rate is not an invariant clock; on clean embeddings it is both clinically meaningful and drug sensitive.

### Evidence for a subcortical interpretation and implications of the equation

The order parameter *ψ* is derived from peripheral and cortical recordings, not from direct brainstem measurements. Converging lines of evidence—a slow 30–50 s spectral component, multi-organ necessity of the embedding, respiratory locking of jumps, and brainstem-targeted pharmacology under dexmedetomidine—make a subcortical interpretation plausible but not proven. Direct subcortical recordings will be required to establish anatomical origin. CAP-based NFLE/RBD contrasts are omitted from the main text after de-clocking.

None of these lines alone establishes a brainstem origin, and several of them rely on the same embedding, so they are not fully independent. Their convergence makes a subcortical interpretation plausible but not proven. Direct subcortical recordings—multi-unit recordings in non-human primates or intraoperative brainstem recordings—will be required to establish the anatomical origin.

### Clinical Interventions Across Orthogonal Axes

From a systems-physiology standpoint, nocturnal sleep offers a natural test of autonomic regulation: with volitional cortical inputs disengaged, somatic homeostasis depends largely on the brainstem central autonomic network, notably the nucleus tractus solitarii (NTS) and rostral ventrolateral medulla (RVLM)[21]. The Landau–Ginzburg description offers a common language for several clinical phenomena that are usually considered separately.

#### Mechanical and pharmacological actions on different axes

In the paired cohort, CPAP relaxed axis A without changing curvature, consistent with relief of the acute respiratory load. The classical GABA_A_ positive allosteric modulator temazepam deepened the well and reduced the bias without changing the subcortical Poisson jump rate, consistent with Kramers escape theory: benzodiazepines may reduce macroscopic awakenings through kinetic confinement rather than by suppressing micro-impulses[22]. Dual orexin receptor antagonists (DORA), by contrast, leave the proportion of slow-wave sleep (N3%) largely unchanged[23]. Within our framework, this predicts an action on the bias axis, lowering *h*, rather than on well depth; given the well-characterized pharmacology of the orexin system[24, 25], this prediction can be tested directly in DORA trials.

## Limitations

Several limitations must be acknowledged. First, for the DISE cohort, because recordings were inherently short, only a subset met the stringent data-density threshold required for stable non-parametric estimation of second-order diffusion terms; nevertheless, primary 1D projection and drift parameters were successfully resolved across all subjects. Second, while our findings align with known noradrenergic and cholinergic neuromodulatory pathways, non-invasive polysomnography cannot directly verify subcortical spike timing; invasive multi-unit recordings in non-human primate models will be needed to directly test the subcortical origin of the jump-diffusion terms. Third, in Sleep-EDFx, missing ECG (cassette) and missing ECG and respiratory channels (telemetry) were zero-padded in the embedding space; because the temazepam trial comes from the telemetry subset, the effect of zero-padding on the order parameter should be quantified, for example by masking these channels in complete four-modality recordings.

## Conclusion

In conclusion, self-supervised multimodal foundation model representations of human sleep converge on a one-dimensional order parameter whose dynamics are well described by a Landau–Ginzburg jump-diffusion equation. This links artificial intelligence representation learning, statistical physics, and clinical sleep medicine, and provides a quantitative framework that complements conventional heuristic scoring with testable descriptions of non-equilibrium state transitions.

## Methods

All analyses operate on continuous polysomnography (PSG) segmented into non-overlapping Δ*t* = 5s tokens and projected into an irreducible one-dimensional macroscopic order parameter *ψ*(*t*). For overnight recordings, early- and late-night windows were defined as the first and last continuous 3.5 h of recording; sessions shorter than 7 h were excluded from overnight comparative analyses. All statistical tests were two-sided at *α* = 0.05 unless a directional hypothesis was pre-specified. To remove a Transformer chunk–position-encoding clock artifact of non-overlapping 5-min inference blocks, embeddings were extracted with a sliding 5-min window at 5-s stride retaining only the center token (Extended Data Fig. 1), which collapsed within-block phase-60 variance (R^2^ 0.998→0.0004) and restored jump–diffusion density closure (R^2^ 0.32→0.80).

## Study Cohorts

Paired CPAP Cohort: Overnight laboratory PSG from adults with obstructive sleep apnea (OSA) studied across paired diagnostic and acute CPAP titration nights (*n* = 14 pairs; 28 nights) at Beijing Tsinghua Changgung Hospital. Inclusion required complete data across four modalities: brain/EEG (BAS), respiratory flow/effort (RESP), electrocardiogram (EKG), and electromyogram (EMG).

DISE Cohort: Drug-induced sleep endoscopy recordings under target-controlled dexmedetomidine infusion (*n* = 143 enrolled; duration filter yielded *n* = 115 nights for center-token NIS+/transfer) from OSA patients visiting Beijing Tsinghua Changgung Hospital. Due to the brief recording length (median = 40min), a subset of *n* = 70 fulfilled the occupancy threshold for stable second-order diffusion and jump estimation (≥40 counts/bin across ≥5 bins). CPS Cohort: Diagnostic PSG from Klinikum Esslingen, *n* = 113 as full cohort; *n* = 84 meeting length and binning criteria for full Kramers–Moyal and jump-diffusion inversion; *n* = 75 with complete AHI, ordinal BMI, and ordinal age for cross-sectional regressions. The cohort is de-identified and lacks gender fields.

Sleep-EDFx: Ambulatory cassette (*n* = 153) and hospital telemetry (*n* = 44) recordings for large-scale *q*^*^ replication (*n* = 197). The telemetry subset contained a double-blind, crossover trial of temazepam versus placebo (*n* = 22 completed pairs). Missing channels (EKG in cassette; EKG/RESP in telemetry) were zero-padded in the embedding space.

### Multimodal Foundation Embeddings and Microscopic Baseline

Continuous waveforms were resampled to *f*_*s*_ = 128Hz and tokenized into 5 s blocks (640 samples/channel). Each token was encoded by the pretrained, frozen SleepFM-Clinical model alongside its preceding 5 min receptive context (60 tokens). Each modality yields a 128 dimensional representation, concatenated at time *t* into a joint embedding *X*(*t*) ∈ ℝ^512^. For historical cohorts lacking specific channels, the corresponding 128-dimensional block was zero-padded. The microscopic baseline state *X*_*t*_ ∈ ℝ^16^ was defined by the leading 16 principal components of *X*(*t*), capturing 60.4% of the cumulative variance in the primary cohort (76.3% and 95.5% for 32 and 128 components).

### Causal Emergence and Order Parameter Identification (NIS+)

To identify macroscopic representations with maximal causal power over microscopic noise, we deployed the NIS+ framework[7]. Bottleneck architectures were trained on normalized information consecutive microscopic pairs (*x*_*t*_,*x*_*t*+Δ*t*_)within-sample for each clinical condition. Effective information eff(*q*) was estimated for candidate latent dimensions *q* ∈ {1,2, 3, 4, 6,8, 12, 16} using the information-sieve approximation. Causal emergence was quantified as Δeff(*q*) = eff(*q*) – eff (16). Across all cohorts—Paired CPAP, CPS, CAP SleepDB, Sleep-EDFx, and DISE, Δeff(*q*) peaked at *q*^*^ = 1. The 1D bottleneck encoder was applied to extract the scalar order parameter, *ψ* (*t*)at 5 s resolution for all subsequent analyses. Biometric trait residualization was evaluated by regressing windowed PCA16 features against sleep stage, AHI, arousal index, and demographics using out-of-fold gradient boosting (GroupKFold by subject). Residual fingerprints were matched across diagnostic and CPAP nights via cosine retrieval, benchmarked against a 500-permutation null.

### Stochastic Differential Equation (SDE) Inversion

The macroscopic order parameter, *ψ* (*t*)was modeled as an overdamped jump-diffusion process under the Itô/Risken convention 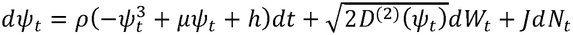 with regularized diffusion *D* ^(2)^ (*ψ*) = max (*ε, d*_0_ *d*_1_*ψ*, d_2_*ψ*^2^) (*ε* = 10^− 12^). The drift fitting using a non-parametric drift 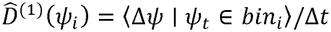 was estimated across *N*_*bins*_ = 28 bins spanning the central 1st–99th percentile range (≥18samples/bin) and mapped to the cubic LG force *D*^(1)^(*ψ*) = *ρ* (− *ψ* ^*3*^ + μ *ψ* + *h*)via weighted least squares 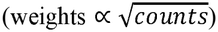. Linear OU drift served as the nested baseline. We also tested non-autonomous time-dependent LG alternatives (TD-LG) incorporating an exponential within-night relaxation clock; this yielded a median timescale of τ= 4.0 h but improved AIC in only 36% of nights (median Δ*NLL* = − 0.0014, leaving autonomous drift favored in 64% of nights). Overnight homeostatic unloading was therefore quantified via paired early-versus late-night half-night potential contrasts rather than an explicit continuous relaxation parameter. Meanwhile, in multiplicative diffusion, the drift residuals δ_*t*_ = Δ*ψ*_*t*_ − *D*^(1)^(*ψ*)_*t*_Δ^*t*^ were binned (16 bins over the 1st–99th percentiles; ≥40 samples/bin across ≥5 valid bins) to estimate the conditional second Kramers–Moyal coefficient 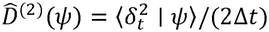, fitted to a quadratic polynomial *d*_o_ *d*_1_*ψ*, d_2_*ψ*^2^. A nested *F*-test evaluated quadratic versus constant diffusion (*H*_o_:*d*_1_ = *d*_2_ = 0). In Jump-Diffusion, standardized residuals 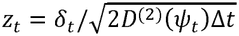 were modeled as a two-component mixture: (1− π)*N(*0,s^2^)+ π *AsymLaplace (b*_*+*_,*b*_*−*_). Discrete jumps were identified at |*z*_*t*_| > 3.0, with arrival intensity 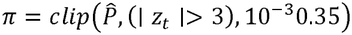. Model selection across nested drift (OU vs. LG) and stochastic noise hierarchies (additive Gaussian, multiplicative Gaussian, multiplicative Student-*t*, and Jump-Diffusion) was evaluated via AIC and out-of-sample negative log-likelihood (NLL) under both cross-subject leave-one-out (LOO) and intra-night temporal (early ↔ late) splits (**Supplemental Table S2**). Overall, the effective potential and global generative closure, was under the Itô/Risken convention for continuous state-dependent diffusion (*J* = 0), the exact stationary log-density potential is 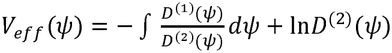. Analytical closure was verified by comparing *V*_*eff*_ (*ψ*)(versus the misspecified Stratonovich-like 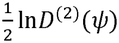entropic term) against log-densities 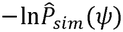 from pure-diffusion (*J* = 0) forward simulations. To test global generative closure of the full jump-diffusion system against real human data, trajectories were forward-simulated from Eq. (3) with empirical (*D*^(1)^, *D*^(2)^, π,*J*) parameters and compared against empirical whole-night log-densities 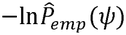 via ordinary least-squares *R*^2^ (contrasting clean center-token trajectories against naive block embeddings).

### Phenotypic Decomposition, Frozen-Ruler Interventions, and Cross-Scale Validation

Overnight homeostatic unloading was quantified by potential barrier height *well_depth =*min(*V*_*peak,L*_,*V*_*peak,R*_*)−V(*ψ_min_) across paired early and late 3.5-h windows, while local bottom curvature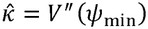 served as a negative-control curvature readout. Rather than drift curvature 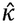, OSA severity in the CPS cohort was tested against the state-dependent multiplicative diffusion coefficients (*d*_0_, *d*_1_,*d*_2_), whitened jump intensity π, and overall trajectory dispersion *y*_*std*_ using Spearman rank correlations. To ensure all intervention and contrast cohorts shared an identical physical coordinate and scaling, embeddings extracted with the sliding-window center-token pipeline from Paired-CPAP, SleepEDFx, and DISE were zero-shot projected through the frozen CPS PCA16 transformation, scaling parameters, and trained 1D NIS+ (*q*^*^ = 1) encoder. In the Paired-CPAP cohort projected onto the frozen CPS ruler, treatment effects (CPAP minus diagnostic night) on baseline diffusion *d*_0_, 30–60 s apnea-band spectral excess, velocity spectral slope *a*_*v*_, jump intensity π (and incremental kurtosis), and late-night (last 3.5-h) well_depth and 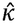 were evaluated using two-sided paired Wilcoxon signed-rank tests. In SleepEDFx, it is projected onto the frozen CPS ruler, paired drug-minus-placebo differences were evaluated via Wilcoxon signed-rank tests, reporting both pre-specified directional one-sided and two-sided *p*-values across continuous well_depth/*d*_0_/apnea-band excess, spectral *a*_*v*_, discrete jump is whitened intensity π and unwhitened absolute jump rate, and clinical awakening *NAW* ≥ 60 s as endpoints, as well as the group-level escape fraction *NAW*/*n*_*jump*_. To test whether individual awakening reductions tracked suppression of jump attempts, within-subject changes Δπ vs. Δ*NAW* were evaluated by Spearman correlation. In DISE cohort, since sessions are substantially shorter than overnight CPS PSGs, each CPS night was randomly subsampled three times into 600-token (50-min) windows; per-night medians of well_depth, 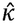, *π, y*_*std*_, *d*_0_, and *d*_2_ across the three subsamples formed the duration-matched CPS baseline. DISE recordings were compared against both CPS-50 min and full-night CPS using two-sided Mann–Whitney *U* tests. To test cross-scale dynamical invariance independently of SleepFM and NIS+, the same Kramers–Moyal jump-diffusion inversion was applied directly to raw cortical EEG beta-band power envelopes 15–30 Hz extracted at ±5s across CPS nights with manual arousal scoring. Subject-specific raw-envelope jump attempt rates *π*_*β*_, effective barrier heights Δ*V*_*β*_ = *V*_*ef,f,β*_(*ψ*_*saddle*_) − *Vef f*,_*β*_(*ψ*_*well*_), and full Kramers escape rates *Pescape*,_*β*_ = *π*_*β*_ exp(−Δ_*β*_/*D*_*β*_) were evaluated against clinical polysomnographic ARI using Pearson correlations and Spearman rank correlations.

### Microstructural Validation and Hypothesis Falsification

Surrogate Testing: Phase-randomized surrogates preserving the exact Fourier amplitude spectrum were generated to confirm that potential well geometry requires non-equilibrium, non-linear temporal sequence structure.

PTT and Autonomic Coincidence: PTT was extracted from synchronized ECG and PPG in CPS. Analysis focused on the late-night EEG-quiet mask. Coincidence between SDE jumps (|*z*| > 3) and clinical autonomic arousals / PTT drops (> 15*ms*) was tested within ±15*s* against Poisson shuffle nulls, supplemented by cross-correlograms (± 60*s*) and transfer entropy.

Rejected Models: (i) Kalman filtering of single-channel PTT failed to track, ψ (*t*) (late-night median *r* = 0.02); (ii) instantaneous fast-slow homeostatic gating 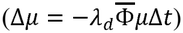 failed to explain variance (*R*^2^ = 0.0082, Δ*AIC* = ±224); (iii) pure demographic noise 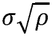 was rejected as it dictates an upward-sloping linear diffusion tensor (*d*_1_ > 0), contradicting empirical quadratic observations (*d*_1_ <0, *d*_2_ > 0).

### Statistical Inference and Software

Within-subject paired comparisons were evaluated via paired *t*-tests (parametric) or Wilcoxon signed-rank tests (non-parametric). Unpaired cohort metrics (e.g., DISE vs. CPS *d*_2_) were assessed using Mann–Whitney *U* tests. Effect sizes are reported as Cohen’s *d*_*z*_ for paired tests and Hedges’ *g* for unpaired contrasts. Associations with clinical covariates were estimated via OLS multiple linear regression. Exploratory comparisons were corrected using the Benjamini–Hochberg False Discovery Rate (FDR; *q*< 0.05). Pipelines were executed in Python (v3.10) using numpy, scipy, pandas, statsmodels, scikit-learn, and torch. SleepFM-Clinical was instantiated using official pretrained weights.

## Data Availability

All data produced in the present study are available upon reasonable request to the authors

## Abbreviations

AASM: American Academy of Sleep Medicine
AHI: apnea–hypopnea index
AIC: Akaike information criterion
ARI: arousal index
BMI: body mass index
CAP: cyclic alternating pattern
CPAP: continuous positive airway pressure
DISE: drug-induced sleep endoscopy
DORA: dual orexin receptor antagonist
ECG/EKG: electrocardiogram
EEG/BAS: electroencephalogram
EMG: electromyogram
JD: jump-diffusion
KM: Kramers–Moyal
LC: locus coeruleus
LG: Landau–Ginzburg
LTF: long-term facilitation
NAW: number of awakenings
NFLE: nocturnal frontal lobe epilepsy
NIS+: Neural Information Squeezer Plus
NLL: negative log-likelihood
NTS: nucleus tractus solitarii
OLS: ordinary least squares
OSA: obstructive sleep apnea
OU: Ornstein–Uhlenbeck
PCA: principal component analysis
PSG: polysomnography
PTT: pulse transit time
RBD: REM sleep behavior disorder
RESP: respiratory effort
RVLM: rostral ventrolateral medulla
SDE: stochastic differential equation
SHE: sleep-related hypermotor epilepsy

## Data Availability Statement

CPS and Sleep-EDFx are publicly available datasets. The Paired CPAP and DISE cohorts are not publicly available under hospital regulations; de-identified data may be requested from the corresponding authors subject to ethics approval. The analysis and simulation source code is available on GitHub once published.

## Ethical Statement

This study was conducted in accordance with the declaration of Helsinki. The hospital cohort was approved by the institutional review board of Beijing Tsinghua Changgung Hospital (IRB No. 26039-0-06), which waived the requirement for informed consent due to the retrospective nature of the analysis.

## Supplemental Material

**Table S1.**
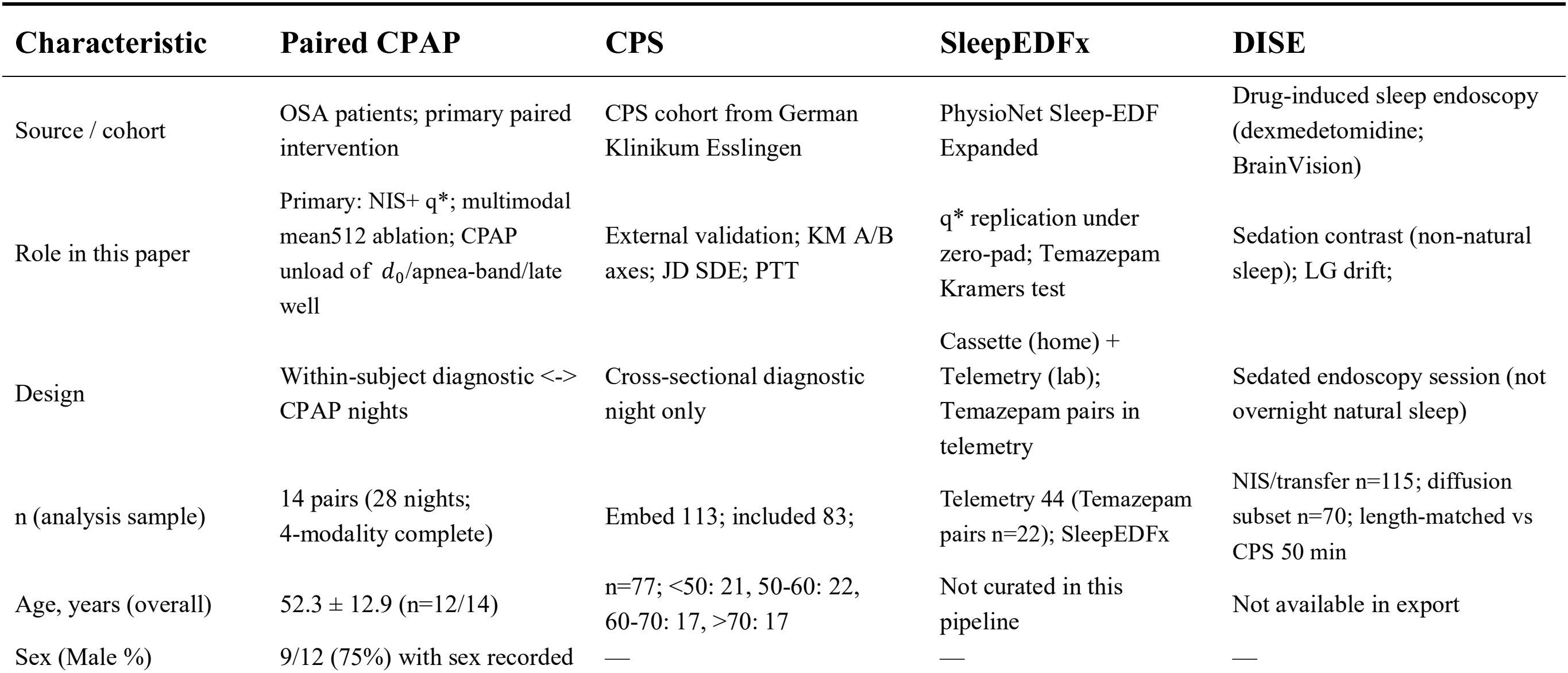
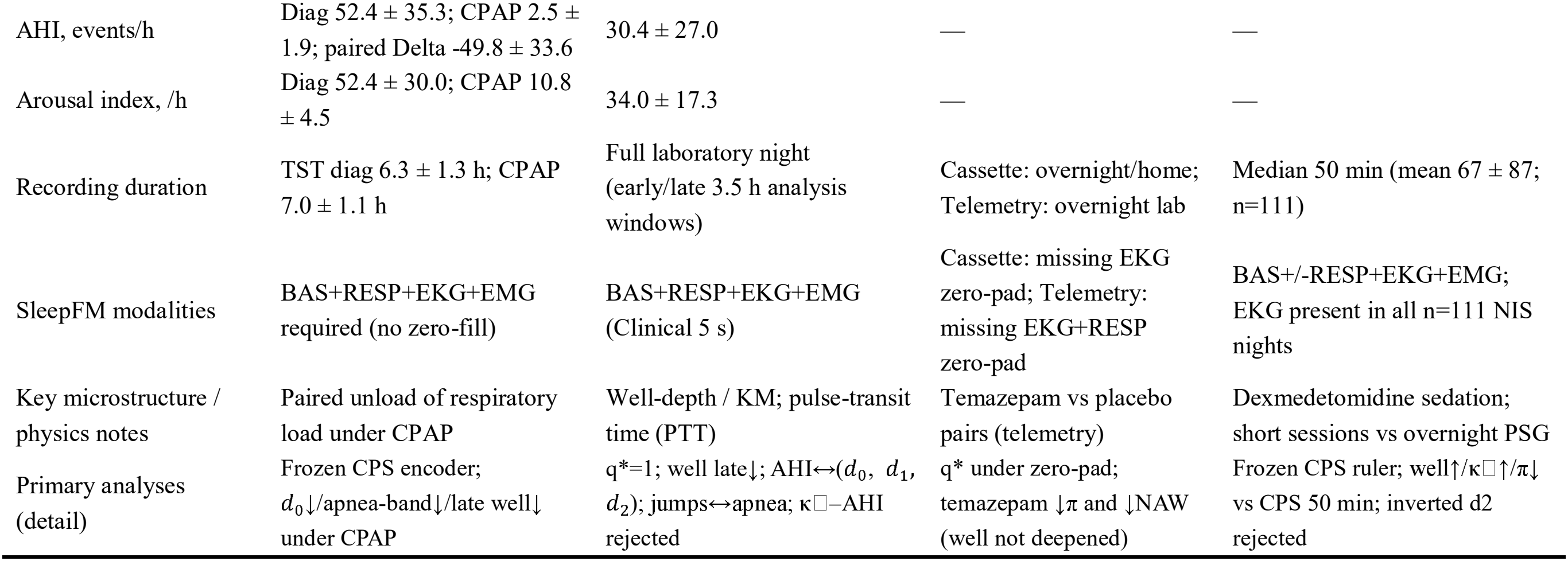
Baseline Characteristics for Cohorts.

| Characteristic | Paired CPAP | CPS | SleepEDFx | DISE |
|---|---|---|---|---|
| Source / cohort | OSA patients; primary paired intervention | CPS cohort from German Klinikum Esslingen | PhysioNet Sleep-EDF Expanded | Drug-induced sleep endoscopy (dexmedetomidine; BrainVision) |
| Role in this paper | Primary: NIS+ q\*; multimodal mean512 ablation; CPAP unload of *d*<sub>0</sub>/apnea-band/late well | External validation; KM A/B axes; JD SDE; PTT | q* replication under zero-pad; Temazepam Kramers test | Sedation contrast (non-natural sleep); LG drift; |
| Design | Within-subject diagnostic <-> CPAP nights | Cross-sectional diagnostic night only | Cassette (home) + Telemetry (lab); Temazepam pairs in telemetry | Sedated endoscopy session (not overnight natural sleep) |
| n (analysis sample) | 14 pairs (28 nights;<br>4-modality complete) | Embed 113; included 83; | Telemetry 44 (Temazepam pairs n=22); SleepEDFx | NIS/transfer n=115; diffusion subset n=70; length-matched vs CPS 50 min |
| Age, years (overall) | 52.3 ± 12.9 (n=12/14) | n=77; <50: 21, 50-60: 22, 60-70: 17, >70: 17 | Not curated in this pipeline | Not available in export |
| Sex (Male %) | 9/12 (75%) with sex recorded | — | — | — |
| AHI, events/h | Diag 52.4 ± 35.3; CPAP 2.5 ± 1.9; paired Delta -49.8 ± 33.6 | 30.4 ± 27.0 | — | — |
| Arousal index, /h | Diag 52.4 ± 30.0; CPAP 10.8 ± 4.5 | 34.0 ± 17.3 | — | — |
| Recording duration | TST diag 6.3 ± 1.3 h; CPAP 7.0 ± 1.1 h | Full laboratory night (early/late 3.5 h analysis windows) | Cassette: overnight/home; Telemetry: overnight lab | Median 50 min (mean 67 ± 87; n=111) |
| SleepFM modalities | BAS+RESP+EKG+EMG required (no zero-fill) | BAS+RESP+EKG+EMG (Clinical 5 s) | Cassette: missing EKG zero-pad; Telemetry: missing EKG+RESP zero-pad | BAS+/-RESP+EKG+EMG; EKG present in all n=111 NIS nights |
| Key microstructure / physics notes | Paired unload of respiratory load under CPAP | Well-depth / KM; pulse-transit time (PTT) | Temazepam vs placebo pairs (telemetry) | Dexmedetomidine sedation; short sessions vs overnight PSG |
| Primary analyses (detail) | Frozen CPS encoder; *d*<sub>0</sub>↓/apnea-band↓/late well↓ under CPAP | q\*=1; well late↓; AHI↔(*d*<sub>0</sub>, *d*<sub>1</sub>, *d*<sub>2</sub>); jumps↔apnea; κ□–AHI rejected | q* under zero-pad; temazepam ↓π and ↓NAW (well not deepened) | Frozen CPS ruler; well↑/κ□↑/π↓ vs CPS 50 min; inverted d2 rejected |

**Table S2.** Out-of-sample model comparison (CPS cohort, mean NLL per 5-s transition)

| **Model Comparison** | **Validation Split** | **Win Rate** | **Median** ΔNLL | **Wilcoxon** *P* |
|---|---|---|---|---|
| **Drift: LG vs. OU** | Temporal (early↔late) | 35.1% (59/168) | -0.0050 | 1 |
|  | Subject LOO | 73.0% (65/89) | +0.0012 | 3.28e-08 |
| **Noise: Mul-Gauss vs. Add-Gauss** | Temporal (early↔late) | 56.9% (95/167) | +0.0069 | 0.0447 |
|  | Subject LOO | 85.4% (76/89) | +0.0244 | 3.36e-08 |
| **Tail: Multiply-*t*(*ν* = 4.88) vs. Multiply-Gauss** | Temporal (early↔late) | 71.9% (120/167) | +0.0452 | 2.28e-16 |
|  | Subject LOO | 55.1% (49/89) | +0.0077 | 2.33e-04 |
| **Jumps: JD vs. Multiply-*t*** | Temporal (early↔late) | 100.0% (167/167) | +2.6256 | <1e-16 |
|  | Subject LOO | 100.0% (89/89) | +2.3737 | 1.28e-16 |
| **Jumps: JD vs. Multiply-Gauss** | Temporal (early↔late) | 100.0% (167/167) | +2.6620 | <1e-16 |
|  | Subject LOO | 100.0% (89/89) | +2.3914 | 1.28e-16 |
|  | Temporal (early↔late) | 35.1% (59/168) | -0.0050 | 1 |

Note: ΔNLL > 0 favors the first-named (candidate) model in each row (LG, Mul-Gauss, Mul-t, or JD). Wilcoxon: one-sided test that ΔNLL > 0. Median LOO NLL hierarchy: additive -4.161, multiplicative Gaussian -4.197, multiplicative Student-t (ν̃=10.03) -4.200, jump–diffusion -6.555. LOO = leave-one-subject-out; Temporal = pooled early↔late half-night splits. Source: clean center-token ψ.

